# Survival in adult inpatients with COVID-19

**DOI:** 10.1101/2020.05.25.20110684

**Authors:** Efrén Murillo-Zamora, Carlos Hernández-Suárez

## Abstract

We conducted a nationwide and retrospective cohort study to assess the survival experience and determining factors in adult inpatients with laboratory-confirmed COVID-19. Data from 5,393 individuals were analyzed using the Kaplan-Meier method and a multivariate Cox proportional hazard regression model was fitted. The 7-day survival was 0.822 and went to 0.482, 0.280, and 0.145 on days 15, 21, and 30 of hospital stay, respectively. In the multiple analysis, factors associated with an increased risk of dying were: male gender, age, longer disease evolution before hospital entry, exposure to mechanical ventilator support, and personal history of chronic noncommunicable diseases (namely obesity, type-2 diabetes mellitus, and chronic kidney disease). To the best of our knowledge, this is the first study analyzing the survival probability in a large subset of Latin-American adults with COVID-19 and our results contribute to achieving a better understanding of disease evolution.

## Background

Worldwide, the coronavirus disease 2019 (COVID-19) by severe acute respiratory syndrome coronavirus 2 (SARS-COV-2) pandemic represents unprecedented health and social crisis. The clinical spectrum of SARS-COV-2 infection is wide and includes asymptomatic contagion, mild upper and unspecific respiratory tract symptoms, and severe viral pneumonia [1]. Most of COVID-19 cases have a good prognosis but a subset of patients develop a critical condition and even die [2].

On May 21, 2020, the observed COVID-19 mortality in Mexico has been high and over 6.5 thousand deaths were registered [3] and, among Latin-American countries, is only overcome by Brazil (nearly 18 thousand deaths) [4]. Published data regarding the clinical course of COVID-19 inpatients is scarce. The computed 14-day survival rate in a study that took place in the city of New York (U.S.), and where 2,773 inpatients were analyzed, was around 50% [5].

The evaluation of clinical outcomes in hospitalized patients with SARS-COV-2 infection may help clinicians and epidemiologists better appreciate the disease evolution, and lead to a more efficient allocation of healthcare resources [6]. This study aimed to assess the survival experience and associated factors in a large cohort of hospitalized adult inpatients with laboratory-confirmed COVID-19.

## Methods

### Study design

We conducted a nationwide and retrospective dynamic cohort study focusing on the survival of hospitalized adult patients with laboratory-confirmed (reverse transcription polymerase chain reaction, qRT-PCR) COVID-19. Eligible subjects were identified from the nominal records of a normative and web-based system for the epidemiological surveillance of viral respiratory diseases, which belongs to the Mexican Institute of Social Security *(IMSS*, the Spanish acronym).

### Population

Individuals aged 18 years or above at acute illness onset and with conclusive evidence of COVID-19 by SARS-COV-2 were potentially eligible. Children and teenagers were not enrolled since current data suggest that severe illness is a rare event among them [7]. Subjects with hospital admission date later than May 5, 2020, were excluded, as well as those with missing clinical or epidemiological data of interest. A total of 341 inpatients were excluded (voluntary hospital discharge, 6.5%; aged under 18 years, 6.7%; referred to another health institution, 33.1%; missing information, 53.7%).

### Data collection

Clinical and epidemiological data of interest were collected from the audited database and included demographic characteristics, illness severity (mild-moderate/severe) [8] at hospital admission, the personal history of chronic non-communicable diseases (no/yes; obesity, arterial hypertension, type 2 diabetes mellitus, asthma, chronic obstructive pulmonary disease, and chronic kidney disease). Dates from illness onset, hospital admission, and discharge (if applicable), as well as the exposure to invasive mechanical ventilation during stay (no/yes), were also obtained from the analyzed surveillance system. The analyzed variables are summarized in Table 1.

**Table 1.** Characteristics of study sample, Mexico 2020

|  | **Died**<br>*n* = 1,735 | **Total**<br>*n* = 5,393 | **Follow-up**<br>(person-days) |
| --- | --- | --- | --- |
| **Gender** |  |  |  |
| Female | 577 (33.3) | 1,961 (36.4) | 17,678 |
| Male | 1,158 (66.7) | 3,432 (63.6) | 30,890 |
| **Age group (years)** |  |  |  |
| 18-29 | 23 (1.3) | 231 (4.3) | 1,744 |
| 30-44 | 212 (12.2) | 1,113 (20.6) | 9,397 |
| 45-59 | 651 (37.6) | 2,082 (38.6) | 18,999 |
| 60 or more | 849 (48.9) | 1,967 (36.5) | 18,428 |
| **Days from symptoms onset to hospitalization** |  |  |  |
| <1 | 681 (39.3) | 2,277 (45.2) | 23,421 |
| 1 to 3 | 299 (17.2) | 909 (16.9) | 7,373 |
| $\geq 4$ | 755 (43.5) | 2,207 (40.9) | 17,774 |
| **Disease severity** |  |  |  |
| Mild-moderate | 234 (13.5) | 1,052 (19.5) | 9,451 |
| Severe | 1,501 (86.5) | 4,341 (80.5) | 39,117 |
| **Invasive mechanical ventilation** |  |  |  |
| No | 1,371 (79.0) | 4,895 (90.8) | 43,773 |
| Yes | 364 (21.0) | 498 (9.2) | 4,795 |
| **Hospital stay (days)** |  |  |  |
| 3 or less | 427 (24.6) | 639 (11.9) | 1,283 |
| 4-6 | 372 (21.4) | 641 (11.9) | 3,215 |
| 7-15 | 720 (41.5) | 3,691 (68.4) | 35,147 |
| 16-30 | 201 (11.6) | 394 (7.3) | 7,917 |
| 31 or more | 15 (0.9) | 28 (0.5) | 1,006 |
| *Personal history of:* |  |  |  |

|  | Died<br>*n* = 1,735 | Total<br>*n* = 5,393 | Follow-up<br>(person-days) |
| --- | --- | --- | --- |
| **Obesity (BMI 30 or higher)** |  |  |  |
| No | 1,277 (73.6) | 4,196 (77.8) | 37,757 |
| Yes | 458 (26.4) | 1,197 (22.2) | 10,811 |
| **Arterial hypertension** |  |  |  |
| No | 927 (53.4) | 3,420 (63.4) | 31,099 |
| Yes | 808 (45.6) | 1,973 (36.6) | 17,469 |
| **Type-2 diabetes mellitus** |  |  |  |
| No | 1,033 (59.5) | 3,716 (68.9) | 34,056 |
| Yes | 702 (40.5) | 1,677 (31.1) | 14,512 |
| **Asthma** |  |  |  |
| No | 1,690 (97.4) | 5,247 (97.3) | 47,263 |
| Yes | 45 (2.6) | 146 (2.7) | 1,305 |
| **COPD** |  |  |  |
| No | 1,612 (92.9) | 5,120 (94.9) | 46,072 |
| Yes | 123 (7.1) | 273 (5.1) | 2,496 |
| **Chronic kidney disease** |  |  |  |
| No | 1,562 (90.0) | 5,094 (94.5) | 46,135 |
| Yes | 173 (10.0) | 299 (5.5) | 2,433 |

Abbreviations: **BMI**, body mass index; COPD, Chronic obstructive pulmonary disease
Note: The absolute and relative (%) frequencies are presented

Medical files from the patients and death certificates represent the primary data source of the surveillance system which data base was employed.

### Outcome

We analyzed the survival time of hospitalized COVID-19 adult patients measured as the time elapsed from the date of hospital entry (starting event) to the date of in-hospital death (final event). The censored variable was defined as the patients who did not present the interest event (did not die) during the follow-up period and the date of hospital discharge was used to compute the time-at risk.

### Laboratory methods

Nasopharyngeal and deep nasal swabs were collected from all analyzed patients in order to perform qRT-PCR (SuperScript™ III Platinum™ One-Step qRT-PCR Kits) analysis.

### Statistical analysis

Summary statistics were computed. The Kaplan–Meier method [9] was employed to estimate the probability of survival from the date of hospital entry. We fitted a Cox proportional hazard regression model to evaluate factors associated with the risk in-hospital death. The assumption of proportional hazard was verified by using a Schoenfeld residual-based test. All analyses were performed by using the Stata software (StataCorp. 2017. Stata Statistical Software: Release 15. College Station, TX: StataCorp LLC.).

### Ethical considerations

This study was approved by the Local Ethics in Health Research Committee (601) of the *IMSS* (R-2020-601-015).

## Results

Data from 5,393 participants (admitted to hospital in a period of 62 days from March 4, to May 5, 2020) were analyzed for a total follow-up of 48,568 person-days. The overall COVID-19 in-hospital lethality rate (*n*= 1,735) was 35.7 per 1,000 person-days. The mean hospital stay (± standard deviation) was 8.4 ± 6.4 vs. 9.3 ± 4.0 days in cases with fatal and nonfatal outcome, respectively *(p<* 0.001).

Table 1 shows the characteristics of participants for selected variables. Most of them were male (63.6%) and 3 out of 4 were aged 45 years or above at hospital admission. Severe illness at entry was documented in 80.5% of participants. In general and as is also shown in Table 1, enrolled patients had a high prevalence of analyzed chronic noncommunicable illnesses.

The Kaplan-Meier survival estimators are presented in Figure 1. A total of 153 deaths were registered within the first day of stay. The survival probabilities of COVID-19 adult inpatients at different periods (1, 3, 7, 15, 21, and 30 days) from hospital admission are summarized in Table 2. The 7-day survival rate was 0.808 (95% CI 0.791-0.824). After 2 weeks from admission, the survival was below 50% (0.482, 95% CI 0.450-0.513).

**Figure 1:**
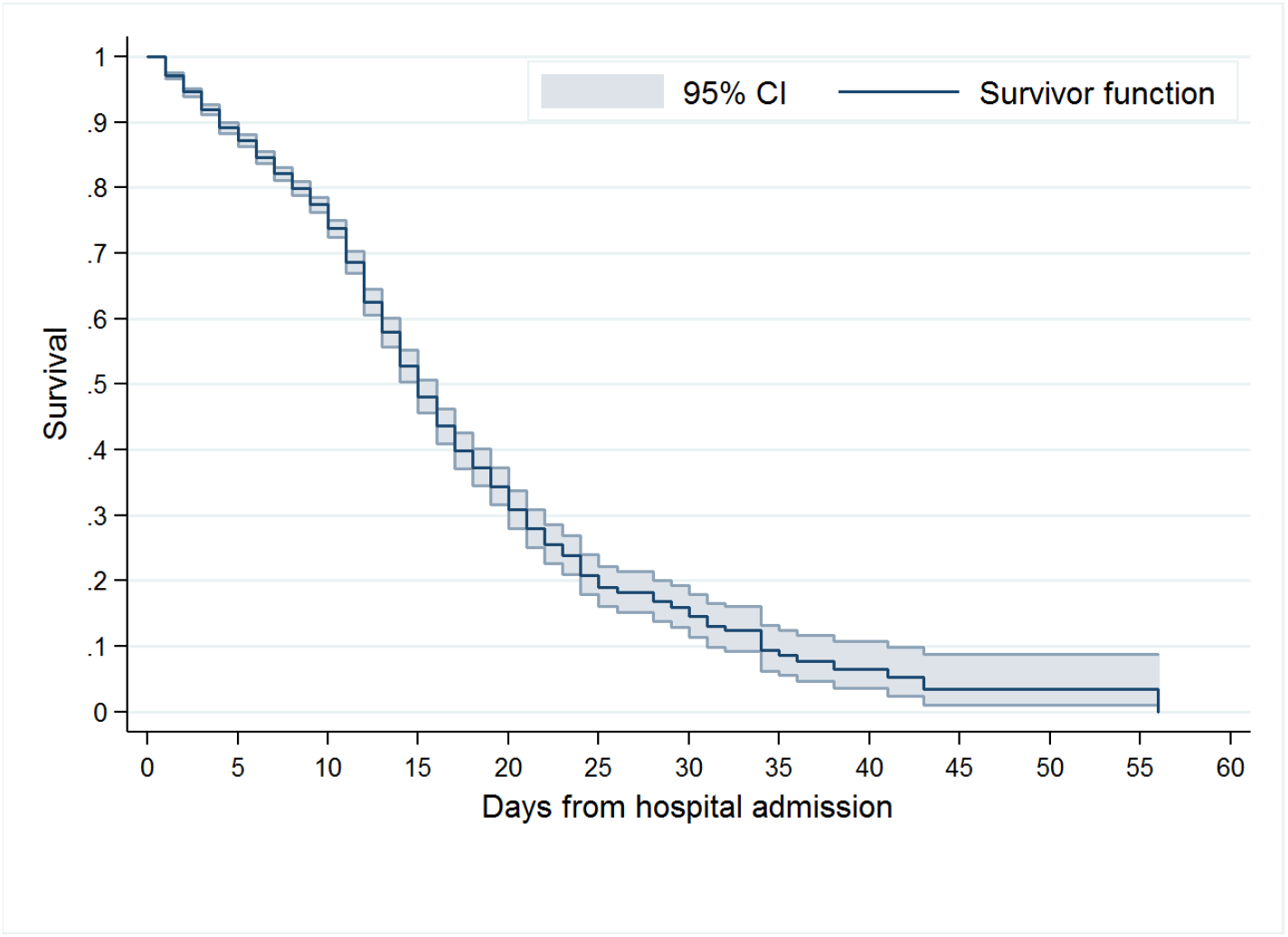
Survival estimators and 95% confidence intervals (CI) in 5,393 adult inpatients with laboratory-confirmed COVID-19, Mexico 2020

**Table 2.** Kaplan Meier survival estimates in adult inpatients with COVID-19, Mexico 2020

| Day | Begin | Deaths | Survival | 95% CI |
| --- | --- | --- | --- | --- |
| 1 | 5,393 | 153 | 0.972 | (0.967-0.976) |
| 3 | 4,982 | 140 | 0.920 | (0.912-0.927) |
| 7 | 4,113 | 122 | 0.822 | (0.811-0.832) |
| 15 | 510 | 45 | 0.482 | (0.456-0.507) |
| 21 | 183 | 17 | 0.280 | (0.250-0.309) |
| 30 | 33 | 2 | 0.145 | (0.114-0.180) |

Abbreviations: **CI**, Confidence interval.

In the multiple model (Table 3), male gender (HR= 1.26, 95% Ci 1-14-1.40) and growing age were associated with an increased risk of in-hospital death. When compared with younger participants (18-29 years), subjects aged 45-59 and 60/above years old, had a 2-fold increase in the risk of dying (45-59 years, HR= 1.99, 95% CI 1.31-3.02; 60 years or above, HR= 2.57, 95% CI 1.69–3.92). Subjects with longer waiting time between symptoms onset and hospital admission also had a lower survival probability ([reference: ¡1 day] 1-3 days, HR= 1.59, 95% CI 1.38-1.82; ≥ 4, HR= 1.68, 95% CI 1.51-1.87), as wells as those with severe manifestations at entry (HR= 1.32, 95% 1.15-1.52).

COVID-19 inpatients requiring ventilatory mechanical support during the stay was also associated with the risk of dying (HR= 1.91, 95%, CI 1.70-2.15). High-risk comorbidities included obesity, type-2 diabetes mellitus, and chronic kidney disease (Table 3).

**Table 3.**
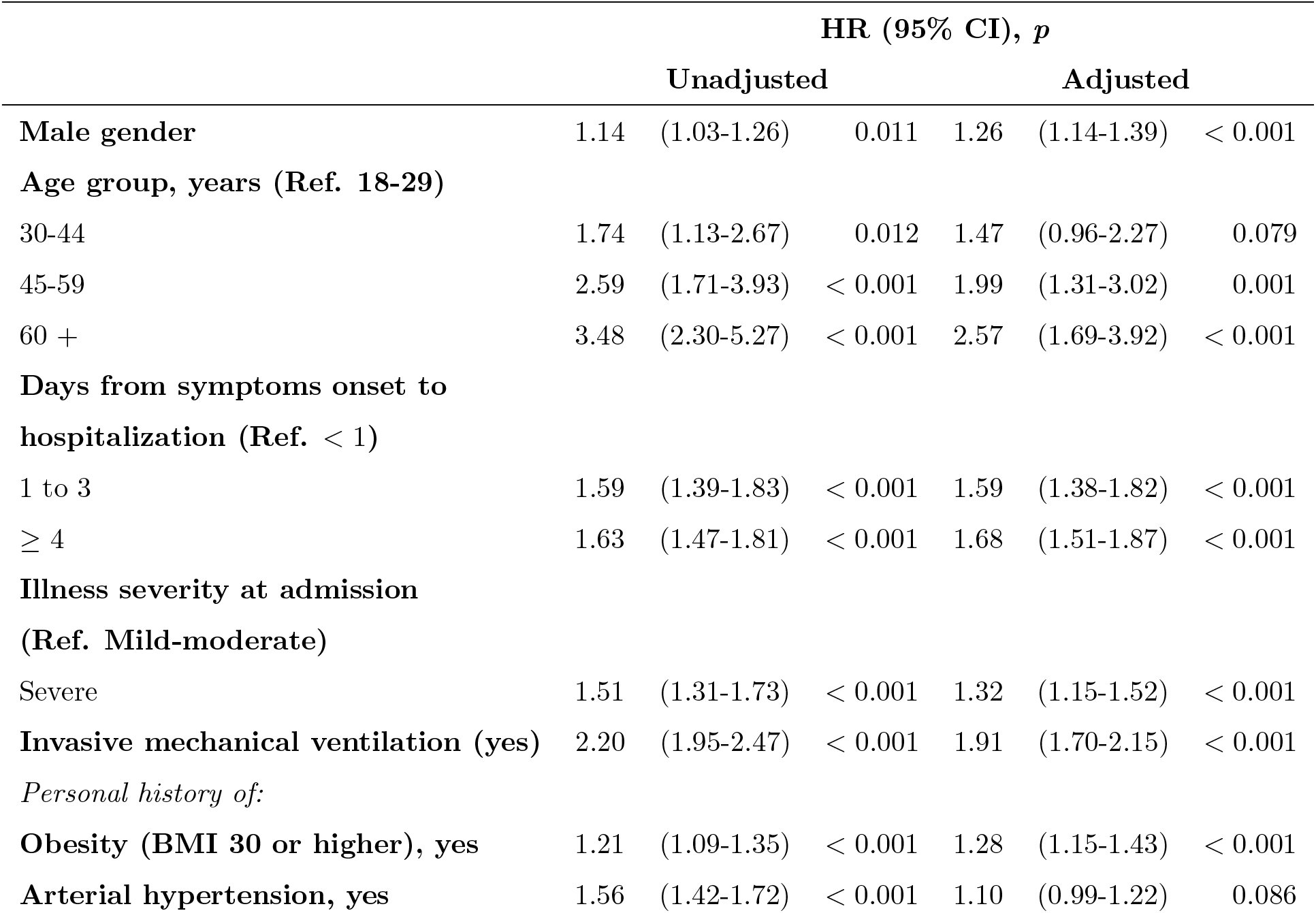
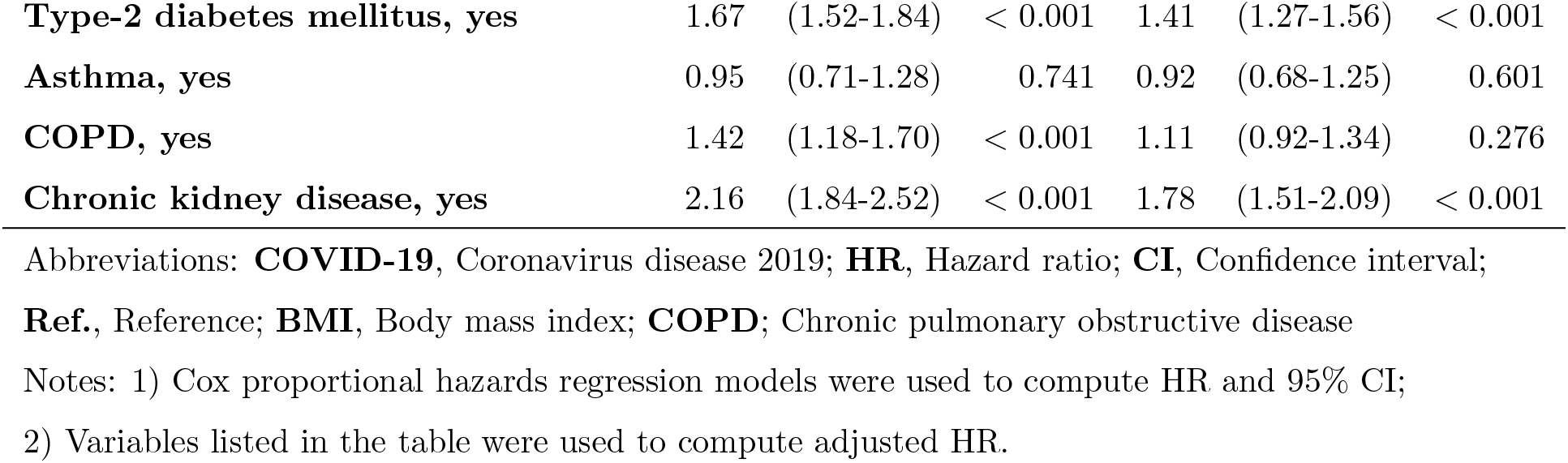
Hazard ratio of dying in COVID-19 adult inpatients, Mexico 2020

## Discussion

The results of this study describe the survival experience of hospitalized adults with COVID-19 and several factors associated with disease outcomes were evaluated. To the best of our knowledge, this is the first study evaluating illness outcomes in a large subset of Latin-American COVID-19 inpatients.

The related burden of SARS-COV-2 in Mexico has been high and obesity and chronic noncommunicable diseases (mainly type-2 diabetes mellitus), both of them showing epidemic characteristics in Mexican adults, may play a role in the observed scenario. Public policy focusing on the prevention of these illnesses has failed and growing trends have been documented [10, 11].

The prevalence of type-2 diabetes mellitus and arterial hypertension in our study sample was significantly higher than national means (diabetes, 31.1% vs. 10.3%, *p<* 0.001; hypertension, 36.6% vs. 18.4%, *p<* 0.001) [12]. These findings were secondary to the inclusion of cases requiring hospitalization; personal history of chronic illness has been associated with a greater risk of severe COVID-19 manifestations and of hospital entry [13].

Gender-related differences have been documented in the severity of SARS-COV-2 symptomatic infection and diseases outcomes. In our study, a shorter survival was observed in males (log-rank test, *p<* 0.001) and, for example, the Kaplan-Meier estimator after one week of hospitalization was 0.840 (95% 0.823-0.856) and 0.810 (95% 0.797-0.824) in women and men, respectively. A protective role of estrogen signaling seems to be involved [14].

Elderly has been consistently associated with death risk among COVID-19 patients and this association is independent from gender and other diseases which frequency also increases with age. In our study, the adjusted HR *per additional year of age* was 1.019 (95% CI 1.015-1.022). Factors determining the age-related risk have not been elucidates but recently published data suggest a role of angiotensin-converting enzyme 2 overexpression together with antibody-dependent enhancement [15].

In our study, longer waiting time between symptoms onset and admission was also associated with survival; participants with longer delay (≥ 4 days), and when compared with those with recent symptoms (<1 day from disease onset to admission), had a 70% increase in the risk of dying (HR= 1.68, 95% 1.51-1.87). Similar findings were described in Hubei, China [16], however the mean elapsed days in our study sample was lower (3.1 vs. 5.7).

Patients requiring mechanical ventilator support during stay had a nearly 2-fold (HR= 1.96, 95%, CI 1.75-2.21) in death risk. This seems to be an effect of the illness severity rather than a cause, since ventilator support was needed in 10.4% vs.4.5% (*p*< 0.001) of severe and mild-moderate cases, respectively. However, and despite the use of these mechanical devices, COVID-19 patients commonly complicate with organ failure or shock [17]. In addition, bacterial coinfections related to invasive therapeutic procedures may play an undetermined role in disease outcomes [18].

The inclusion of only laboratory-positive cases, together with the large sample size and national representativeness, are major strengths of this study. However, potential limitations must be cited. First, we were unable to assess a gradient between body mass and survival functions, since anthropometric registers are not collected by the audited epidemiological surveillance system. Instead, obesity data is collected as a dichotomous variable. And second, no biomarkers data were available and which may have improved the accuracy of built models. Among others, a prognostic value of B-type natriuretic peptide and creatine kinase-MB has been documented recently [19].

## Conclusion

The COVID-19 pandemic-related mortality in Mexico has been high. The survival experience of hospitalized adults was documented in this nation-wide study and factors determining the illness outcome were assessed. Since obese and type 2 diabetes mellitus patients had a poor prognosis, our results highlight the major relevance of public health policies and interventions focusing on their prevention in the analyzed population.

## Data Availability

The data that support the findings of this study are available on request from the corresponding author.

